# Effectiveness of mRNA COVID-19 vaccines against symptomatic SARS-CoV-2 infections during the Delta variant epidemic in Japan: Vaccine Effectiveness Real-time Surveillance for SARS-CoV-2 (VERSUS)

**DOI:** 10.1101/2022.01.17.22269394

**Authors:** Haruka Maeda, Nobuo Saito, Ataru Igarashi, Masayuki Ishida, Kazuya Suami, Ai Yagiuchi, Yuya Kimura, Masaru Komino, Hiromi Arai, Toru Morikawa, Iori Motohashi, Rei Miyazawa, Tetsu Moriyama, Hiroshi Kamura, Mayumi Terada, Osamu Kuwamitsu, Tomoichiro Hayakawa, Eiichiro Sando, Yasuji Ohara, Osamu Teshigawara, Motoi Suzuki, Konosuke Morimoto

## Abstract

**Background:** Although high vaccine effectiveness of messenger RNA (mRNA) coronavirus disease 2019 (COVID-19) vaccines was reported in studies in several countries, data is limited from Asian countries, especially against the Delta (B.1.617.2) variant.

**Methods:** We conducted a multicenter test-negative case-control study in patients aged ≥16 visiting hospitals or clinics with signs or symptoms consistent with COVID-19 from July 1 to September 30, 2021, when the Delta variant was dominant (≥90% of severe acute respiratory syndrome coronavirus 2 [SARS-CoV-2] infections) nationwide in Japan. Vaccine effectiveness of BNT162b2 or mRNA-1273 against symptomatic SARS-CoV-2 infections was evaluated. Waning immunity among patients aged 16 to 64 was also assessed.

**Results:** We enrolled 1936 patients, including 396 test-positive cases and 1540 test-negative controls for SARS-CoV-2. The median age was 49 years, 53.4% were male, and 34.0% had underlying medical conditions. Full vaccination (receiving two doses ≥14 days before symptom onset) was received by 6.6% of cases and 38.8% of controls. Vaccine effectiveness of full vaccination against symptomatic SARS-CoV-2 infections was 88.7% (95% confidence interval [CI], 78.8–93.9) among patients aged 16 to 64 and 90.3% (95% CI, 73.6–96.4) among patients aged ≥65. Among patients aged 16 to 64, vaccine effectiveness within one to three months after full vaccination was 91.8% (95% CI, 80.3–96.6), and was 86.4% (95% CI, 56.9–95.7) within four to six months.

**Conclusions:** mRNA COVID-19 vaccines had high effectiveness against symptomatic SARS-CoV-2 infections in Japan during July 1 to September 30, 2021, when the Delta variant was dominant nationwide.

## Introduction

Since December 2019, the severe acute respiratory syndrome coronavirus 2 (SARS-CoV-2) has spread globally, including Japan, and has significantly impacted health, livelihoods, and economics. To counter the coronavirus disease 2019 (COVID-19) pandemic, COVID-19 vaccines were developed and distributed globally. Clinical trials of COVID-19 vaccines found high vaccine efficacy [1-3], and observational studies evaluated vaccine effectiveness in several countries [4-6]. However, data on vaccine effectiveness of messenger RNA (mRNA) COVID-19 vaccines, especially against the Delta (B.1.617.2) variant, from Asian countries is limited.

In February 2021, the Japanese government initiated a national COVID-19 vaccination campaign (Supplementary Figure 1). It is crucial to assess COVID-19 vaccine effectiveness domestically when evaluating the national policy and, going forward, determining the optimal vaccination policy. Vaccine effectiveness has been estimated to attenuate due to the emergence of new variants [7]. Accordingly, we started surveillance activity from July 1, 2021 to monitor vaccine effectiveness of COVID-19 vaccines in Japan, named Vaccine Effectiveness Real-time Surveillance for SARS-CoV-2 (VERSUS). In this study, we evaluated vaccine effectiveness of mRNA COVID-19 vaccines, BNT165b2 and mRNA-1273, against symptomatic SARS-CoV-2 infections during the Delta variant epidemic in Japan using data registered for our surveillance.

## Methods

### Design

We conducted a prospective test-negative case-control study in patients visiting hospitals or clinics with signs or symptoms compatible with COVID-19 [8, 9]. The case group included individuals having signs or symptoms compatible with COVID-19 and positive test results of SARS-CoV-2. The following test methods were included, which have high sensitivity and specificity and are commonly used for diagnosis in Japan as well as globally [10, 11]: nucleic acid amplification tests including polymerase chain reaction (PCR), loop-medical isothermal amplification (LAMP) [12], and nicking endonuclease amplification reaction [13]; and antigen quantification tests [14, 15]. Individuals having signs or symptoms compatible with COVID-19 but negative test results of SARS-CoV-2 were included in the control group.

### Setting

This study enrolled individuals visiting medical institutes from July 1 through September 30, 2021, at nine hospitals and four clinics in nine prefectures on four main islands in Japan. During this study period, Japan experienced a fifth epidemic wave due to the Delta variant, starting late in June 2021 (Supplementary Figure 2) [16, 17].

Before the fifth wave in Japan, the Japanese government approved and introduced COVID-19 vaccines [18] (Supplementary Figure 1). The market approval of BNT162b2 was done on February 14, 2021 for aged ≥16 years and expanded for aged 12 to 15 years on May 31, 2021. Two additional vaccines, mRNA-1273 and AZD1222, were approved for aged ≥18 years on May 21, 2021. For mRNA-1273 approval was expanded for aged 12 to 17 years on July 26, 2021. The government decided to publicly fund COVID vaccinations and set up a prioritization strategy, with a design based in part on procurement issues. Healthcare workers were the first to be vaccinated, beginning on February 17, 2021, followed by priority vaccination of older adults aged ≥65 years started on April 12, 2021. For younger citizens, 12 to 64 years of age, a vaccination program was started from June 2021, with priority given to those with underlying medical conditions. All were vaccinated with BNT162b2 until May 24. The first administration of mRNA-1273 for older adults aged ≥65 years began on May 24, 2021, followed by vaccination of aged ≥18 years on June 17, 2021, and aged ≥12 years on August 2, 2021. Administration of AZD1222 became optional for aged ≥40 years from late August. By September 30, 2021, 66% of the Japan population had received at least one dose and 57% received two doses; among people aged ≥65 years, more than 90% had received two doses (Supplementary Figure 2) [19].

### Participants

Patients aged ≥16 years were included who had signs or symptoms compatible with COVID-19 (specifically, one or more of the following: fever [≥37.5°C], cough, fatigue, shortness of breath, myalgia, sore throat, nasal congestion, headache, taste disorder, or olfactory dysfunction) [20, 21] and were tested for SARS-CoV-2. We excluded cases with episodes tested 15 days or more after symptom onset or cases with undocumented symptom onset dates, because of the inaccuracy of test results [22]. When individuals had multiple episodes, we used the following rules for exclusion: 1) episodes with negative test results within seven days after a previous negative result; 2) episodes with multiple negative test results and identical symptom onset date; 3) episodes with negative test results within three weeks prior to a positive test result, or episodes occurring after a positive test result, due to the possibility of false-negatives; 4) for multiple positive episodes during the study period, we included only the first episode; and 5) we included a maximum of three negative test results for each individual.

### Data collection

Attending physicians in each medical institute identified eligible patients. Demographic and clinical information was collected from medical charts and recorded on an electronic database using REDCap [23].

We collected demographic and clinical information, including age, sex, place of residence, presence of underlying medical conditions (i.e. chronic heart disease, chronic respiratory disease, obesity [body mass index≥30 kg/m^2^], malignancy [including solid or haematological malignancy], diabetes mellitus, chronic kidney disease, receiving dialysis, liver cirrhosis, use of immunosuppressive medicines, or pregnancy), smoking history, history of contact with COVID-19 patients, healthcare employment status, clinical symptoms, and COVID-19 vaccination histories.

### Classification of vaccination status

We obtained vaccination histories (i.e., vaccination date of administration, type of vaccine product, and vaccination frequency) through interviews with patients or their family members. COVID-19 vaccines are administered as two-dose series. Vaccination status was classified into six categories based on the number of vaccine doses received before symptom onset and the number of days between the last vaccination and symptom onset date; specifically, 1) unvaccinated where individuals had received no vaccine dose before symptom onset; 2) first vaccine dose within 13 days before symptom onset; 3) partially vaccinated where individuals received one dose ≥14 days before symptom onset; 4) second vaccine dose within 13 days before symptom onset; 5) fully vaccinated where individuals received two doses ≥14 days before symptom onset; and 6) unknown vaccination status where vaccination histories were not documented. For patients whose precise vaccination date was not documented (for example, only the month of the vaccination was documented), the midpoint between the two possible dates was assumed to be the vaccination date. Additionally, those for whom only the number of vaccine doses were recorded were included in either the partially or fully vaccinated groups depending on the number of vaccine doses.

### Statistical analysis

The odds ratio (OR) was calculated by comparing the odds of antecedent COVID-19 vaccination in test-positive versus test-negative patients. A mixed-effects logistic regression model was used to calculate adjusted ORs. Age, sex, presence of underlying medical conditions, calendar weeks, and history of contact with COVID-19 patients were applied as the fixed effects, and study sites as the random effect to the logistic model. Vaccine effectiveness was defined as one minus adjusted ORs, expressed as a percentage [8, 9].

Vaccine effectiveness estimates were calculated for full vaccinated versus unvaccinated and for partially vaccinated versus unvaccinated. We analyzed vaccine effectiveness separately in patients aged 16 to 64 years and in patients aged ≥65 years, taking into consideration the possibility of confounders due to the priority vaccination strategy for patients aged ≥65 years. For BNT162b2 or mRNA-1273 analysis, we pooled patients who received either BNT162b2 or mRNA-1273mRNA COVID-19 vaccines. We also performed analyses on each vaccine product separately. We excluded the episodes with undocumented vaccine products from the analysis of each vaccine product. Additionally, to assess the extent of waning immunity of mRNA COVID-19 vaccines against the Delta variant in Japan, we evaluated vaccine effectiveness separately between two groups: episodes within one to three months and episodes within four to six months after full vaccination status (14 days after the second vaccine receipt) among patients aged 16 to 64 years. We also conducted subgroup analyses by sex or presence of underlying medical conditions.

Several sensitivity analyses were performed to strengthen our results since some of the vaccination histories were vacant: when the precise vaccination date was uncertain, we set the vaccination date as the most recent possible date (scenario A) or the earliest possible date (scenario B) from symptom onset; we excluded patients with no information of vaccination dates (scenario C); and multiple imputations were performed for those with undocumented vaccination histories (scenario D). All analysis was performed using Stata version 16.0 (Stata Corp., College Station, Texas, USA)

### Ethics

This study was approved by the Institutional Review Board (IRB) at the Institute of Tropical Medicine, Nagasaki University (approval no. 210225257) and the study sites. For the study sites without IRBs, this study was collectively reviewed by the Institute of Tropical Medicine IRB, Nagasaki University. The study was conducted following the Ministry of Health, Labour and Welfare Ethical Guidelines for Medical and Biological Research Involving Human Subjects 2021 [24].

## Results

### Participants

Between July 1, 2021, and September 30, 2021, 2082 episodes with signs or symptoms consistent with COVID-19 and evidence of tests for SARS-CoV-2 were registered in our surveillance. After excluding 75 episodes with tests ≥15 days after symptom onset, 43 episodes with undocumented symptom onset dates, and 28 episodes with multiple test occasions, we included 1936 patients (396 test-positive cases, 1540 test-negative controls) (Figure 1) enrolled from 13 medical facilities (Supplementary Figure 3). Overall, the median age was 49 years (interquartile range, 30–72), 1033 (53.4%) were male, 659 (34.0%) had one or more underlying medical conditions (Table 1). Test-positive cases were more likely to be male, younger, have histories of contact with COVID-19 patients, and less likely to have underlying medical conditions (p-value <0.001). Thirteen (3.3%) cases and 89 (5.8%) controls had received patrial COVID-19 vaccination, 26 (6.6%) cases and 597 (38.8%) controls had received full COVID-19 vaccination, and 290 (73.2%) cases and 523 (34.0%) controls were unvaccinated. Among vaccinated patients, 676 (69.6%) received BNT162b2 and 140 (14.4%) mRNA-1273 but the vaccine products of 155 (16.0%) individuals were unknown; and 86.9% of fully vaccinated individuals had completed full vaccination within one to three months before symptom onset.

**Figure 1.**
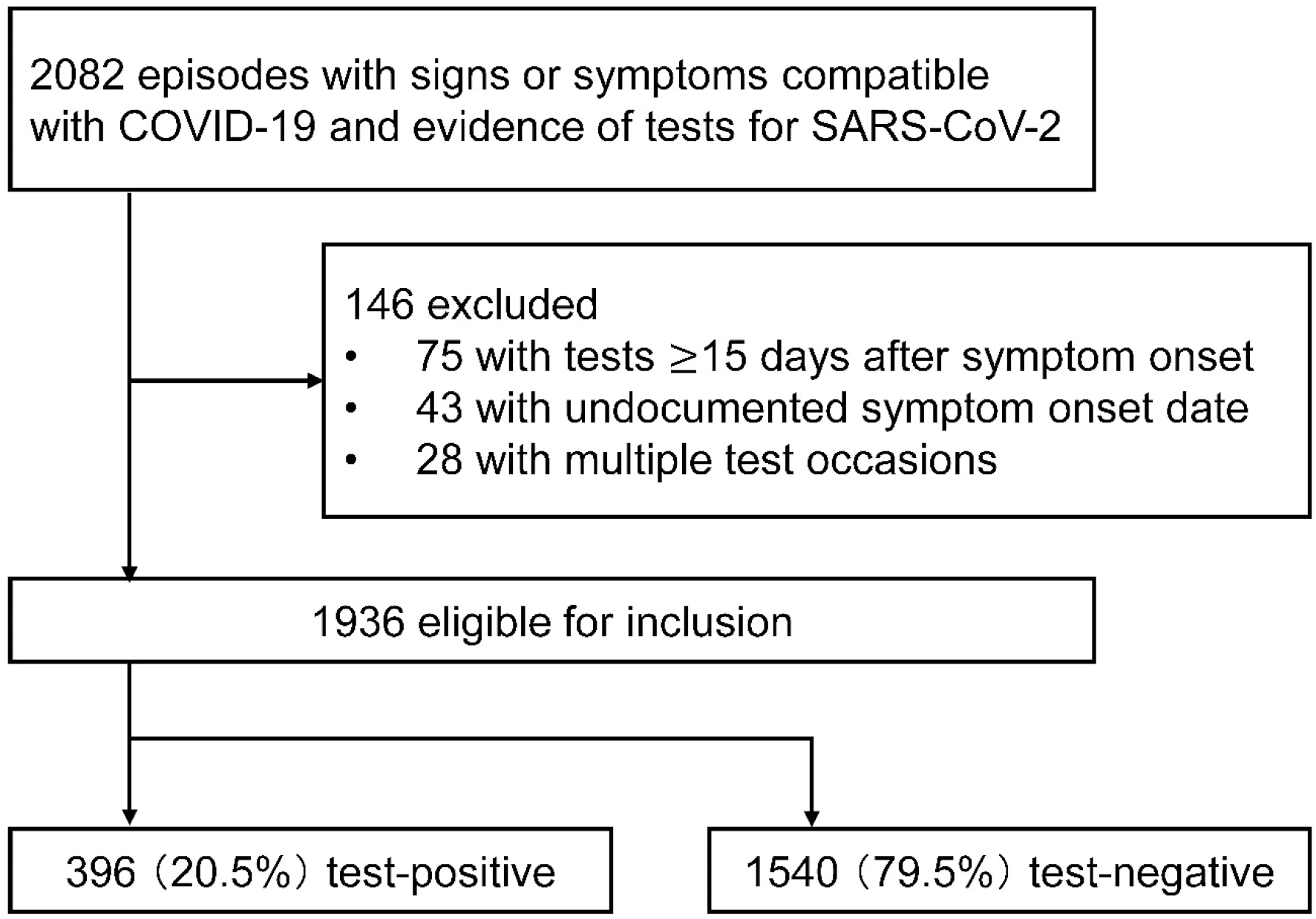
Patients aged ≥16 years were enrolled in this study who had signs or symptoms compatible with COVID-19 (specifically, one or more of the following: fever (≥37.5°C), cough, fatigue, shortness of breath, myalgia, sore throat, nasal congestion, headache, taste disorder, or olfactory dysfunction) and were tested for SARS-CoV-2 in a medical institute in Japan between July 1, 2021 and September 30, 2021. Abbreviations: COVID-19, coronavirus disease 2019; SARS-CoV-2, severe acute respiratory syndrome coronavirus 2.

**Table 1:** Demographics and characteristics of test-positive cases and test-negative controls: VERSUS study, Japan, July 1 to September 30, 2021

|  | **Total**<br>**(n=1936)** | **Test-**<br>**positive**<br>**case**<br>**(n=396)** | **Test-**<br>**negative**<br>**control**<br>**(n=1540)** |
| --- | --- | --- | --- |
| Median age (IQR), year | 49 (30–72) | 35 (26–50) | 55 (32–76) |
| Age category in years, no. (%) |  |  |  |
| 16–29 | 475 (24.5) | 148 (37.4) | 327 (21.2) |
| 30–39 | 278 (14.4) | 78 (19.7) | 200 (13.0) |
| 40–49 | 239 (12.4) | 68 (17.2) | 171 (11.1) |
| 50–59 | 201 (10.4) | 64 (16.2) | 137 (8.9) |
| 60–69 | 195 (10.1) | 12 (3.0) | 183 (11.9) |
| 70–79 | 220 (11.4) | 12 (3.0) | 208 (13.5) |
| 80–89 | 242 (12.5) | 11 (2.8) | 231 (15.0) |
| ≥90 | 86 (4.4) | 3 (0.8) | 83 (5.4) |
| Male sex, no. (%) | 1033 (53.4) | 246 (62.1) | 787 (51.1) |
| Living at home, no. (%) | 1767 (91.3) | 385 (97.2) | 1382 (89.7) |
| Living at a long-term care facility, no. (%) | 121 (6.3) | 1 (0.3) | 120 (7.8) |
| Underlying medical conditions, no. (%) |  |  |  |
| Any | 659 (34.0) | 71 (17.9) | 588 (38.2) |
| Chronic cardiovascular disease | 166 (8.6) | 13 (3.3) | 153 (9.9) |
| Chronic respiratory disease | 182 (9.4) | 11 (2.8) | 171 (11.1) |
| Obesity | 92 (4.7) | 24 (6.1) | 68 (4.4) |
| Malignancy | 148 (7.6) | 8 (2.0) | 140 (9.1) |
| Diabetes mellitus | 181 (9.3) | 20 (5.1) | 161 (10.4) |
| Chronic kidney disease | 76 (3.9) | 3 (0.8) | 73 (4.7) |
| Dialysis | 21 (1.1) | 1 (0.3) | 20 (1.3) |
| Liver cirrhosis | 6 (0.3) | 0 | 6 (0.4) |
| Immunocompromising therapy | 46 (2.4) | 4 (1.0) | 43 (2.7) |
| Pregnancy | 5 (0.3) | 2 (0.5) | 3 (0.2) |
| Smoking history, no. (%) | 624 (32.2) | 143 (36.1) | 481 (31.2) |
| Healthcare workers, no. (%) | 108 (5.6) | 7 (1.8) | 101 (6.6) |
| History of contact with a COVID-19 patient, no. (%) | 217 (11.2) | 131 (33.1) | 86 (5.6) |
| COVID-19 vaccination status<sup>a</sup>, no. (%) |  |  |  |
| Unvaccinated | 813 (42.0) | 290 (73.2) | 523 (34.0) |
| First vaccine dose within 0–13 days | 125 (6.4) | 29 (7.3) | 96 (6.2) |
| Partially vaccinated<br>(first vaccine dose after $\geq 14$ days) | 102 (5.3) | 13 (3.3) | 89 (5.8) |
| Second vaccine doses within 0–13 days | 121 (6.3) | 2 (0.5) | 119 (7.7) |
| Fully vaccinated<br>(second vaccine doses after $\geq 14$ days) | 623 (32.2) | 26 (6.6) | 597 (38.8) |
| Unknown vaccination status | 152 (7.9) | 36 (9.1) | 116 (7.5) |
| Among vaccinated patients, vaccine product received, no (%) |  |  |  |
| BNT162b2 | 676 (69.6) | 42 (60.0) | 634 (71.4) |
| mRNA-1273 | 140 (14.4) | 14 (20.0) | 126 (14.2) |
| Unknown | 155 (16.0) | 14 (20.0) | 141 (15.9) |
| Among fully vaccinated patients with documented vaccination date, months after full vaccination<sup>b</sup>, no (%) |  |  |  |
| One to three months | 458 (86.9) | 19 (82.6) | 439 (87.1) |
| Four to six months | 69 (13.1) | 4 (17.4) | 65 (12.9) |

Abbreviations: SARS-CoV-2, severe acute respiratory syndrome coronavirus 2; VERSUS, Vaccine Effectiveness Real-time Surveillance for SARS-CoV-2; IQR, interquartile range; COVID-19, coronavirus disease 2019.
<sup>a</sup>Vaccination status was classified based on the number of COVID-19 vaccine doses received before symptom onset. Unvaccinated individuals received no COVID-19 vaccine dose before symptom onset. Partially vaccinated individuals received one dose of COVID-19 vaccine $\geq 14$ days before symptom onset. Fully vaccinated individuals received two doses of COVID-19 vaccines $\geq 14$ days before symptom onset.
<sup>b</sup>Excluded those with no information of vaccination date: three test-positive cases and 93 test-negative controls were excluded.

### Vaccine effectiveness

Vaccine effectiveness of mRNA COVID-19 vaccines against symptomatic SARS-CoV-2 infections are shown in Figure 2. For BNT162b2 or mRNA-1273 analysis among patients aged 16 to 64 years, vaccine effectiveness for full vaccination was 88.7% (95% confidence interval [CI], 78.8–93.9), and 54.3% (95% CI, 8.4–77.2) for partial vaccination. Point estimates were higher for mRNA-1273 (96.6%; 95% CI, 72.8–99.6) than for BNT162b2 (86.7%; 95% CI, 73.5–93.3); however, there was no statistically significant difference (p-value=0.877). Among patients aged ≥65 years, vaccine effectiveness for full vaccination was similar to aged 16 to 64 years: 90.3% (95% CI, 73.6–96.4) for BNT162b2 or mRNA-1273 analysis, and 85.8% (95% CI, 59.4–95.0) for BNT162b2 analysis. Vaccine effectiveness for partial vaccination or mRNA-1273 among patients aged ≥65 years was not evaluated due to the small sample size.

**Figure 2.**
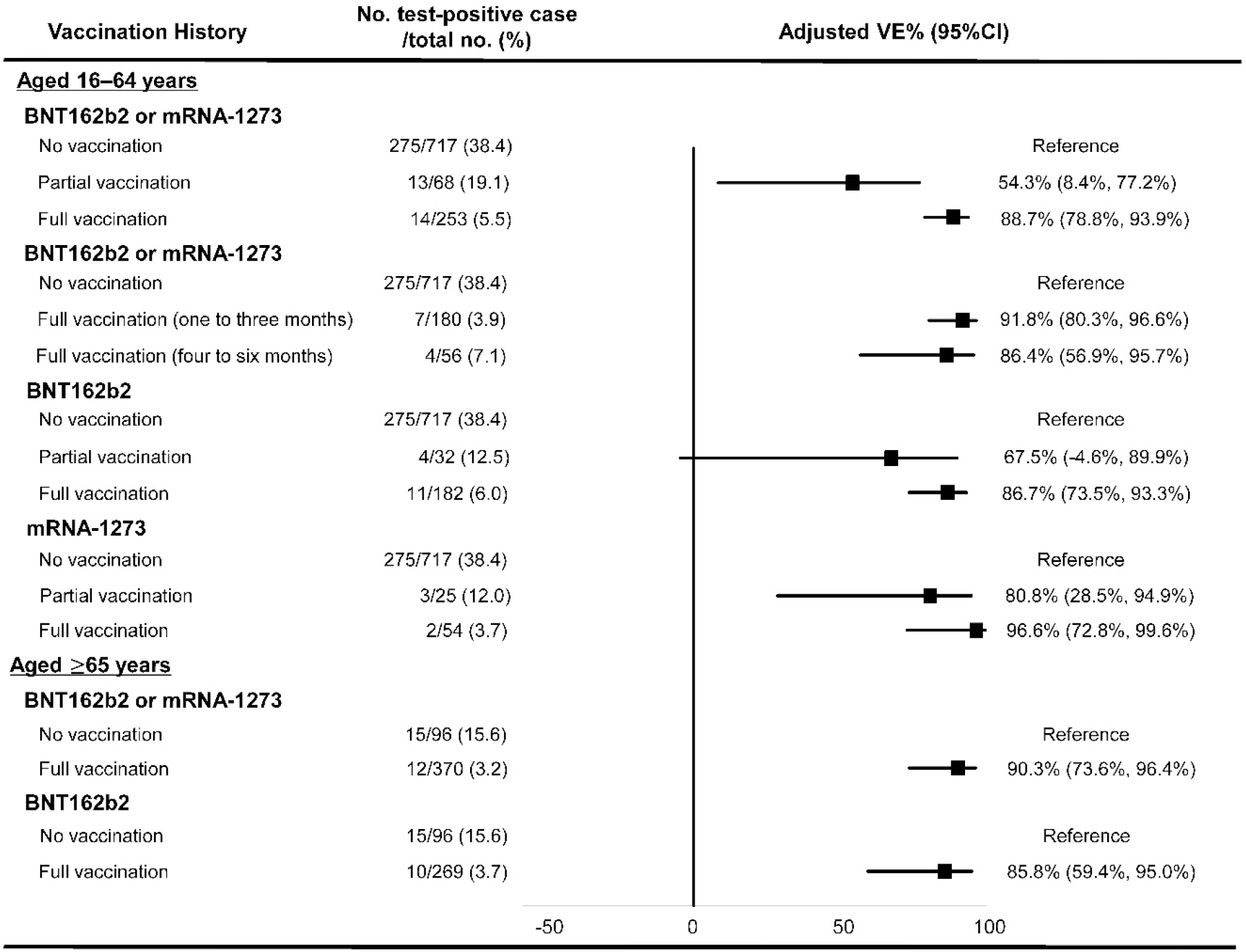
Vaccine effectiveness of messenger RNA COVID-19 vaccines against symptomatic SARS-CoV-2 infections among individuals aged 16 to 64 years and aged ≥65 years, VERSUS study, Japan, July 1–September 30, 2021. The analysis included test-positive cases who had signs or symptoms comparable with COVID-19 and tested positive for SARS-CoV-2, and test-negative controls who had signs or symptoms comparable with COVID-19 and were tested negative for SARS-CoV-2. Vaccine effectiveness were adjusted for age, sex, presence of underlying medical conditions, calendar week, history of contact with COVID-19 patients, and study site. Vaccination status was classified into three statuses based on the number of vaccine doses received before symptom onset and the number of days between the last vaccination date and symptom onset; no vaccination where individuals had received no vaccine dose before symptom onset; partial vaccination where individuals received one dose ≥14 days before symptom onset; and full vaccination where individuals who received two doses ≥14 days before symptom onset. Abbreviations: VE, vaccine effectiveness; COVID-19, coronavirus disease 2019; SARS-CoV-2, severe acute respiratory syndrome coronavirus 2; VERSUS, Vaccine Effectiveness Real-time Surveillance for SARS-CoV-2.

The extent of waning immunity of mRNA COVID-19 vaccines against symptomatic SARS-CoV-2 infections was analyzed among patients aged 16 to 64 years. Vaccine effectiveness for patients within one to three months after full vaccination was 91.8% (95% CI, 80.3–96.6), and 86.4% (95% CI, 56.9–95.7) for patients within four to six months (Figure 2).

In a subgroup analysis by sex, a point estimate was higher for women (92.9%; 95% CI, 81.0–97.4) than for men (83.5%; 95% CI, 62.3–92.8) among patients aged 16 to 64 years, and higher for men (94.1%; 95% CI, 72.5–98.8) than women (88.7%; 95% CI, 49.2–97.5) among patients aged ≥65 years, while the 95% CI overlapped in both age groups (Figure 3). In a subgroup analysis by the presence of underlying medical conditions, vaccine effectiveness was similar for both age groups. The results of sensitivity analysis are shown in Supplementary Table 1 and are similar to the primary analysis.

**Figure 3.**
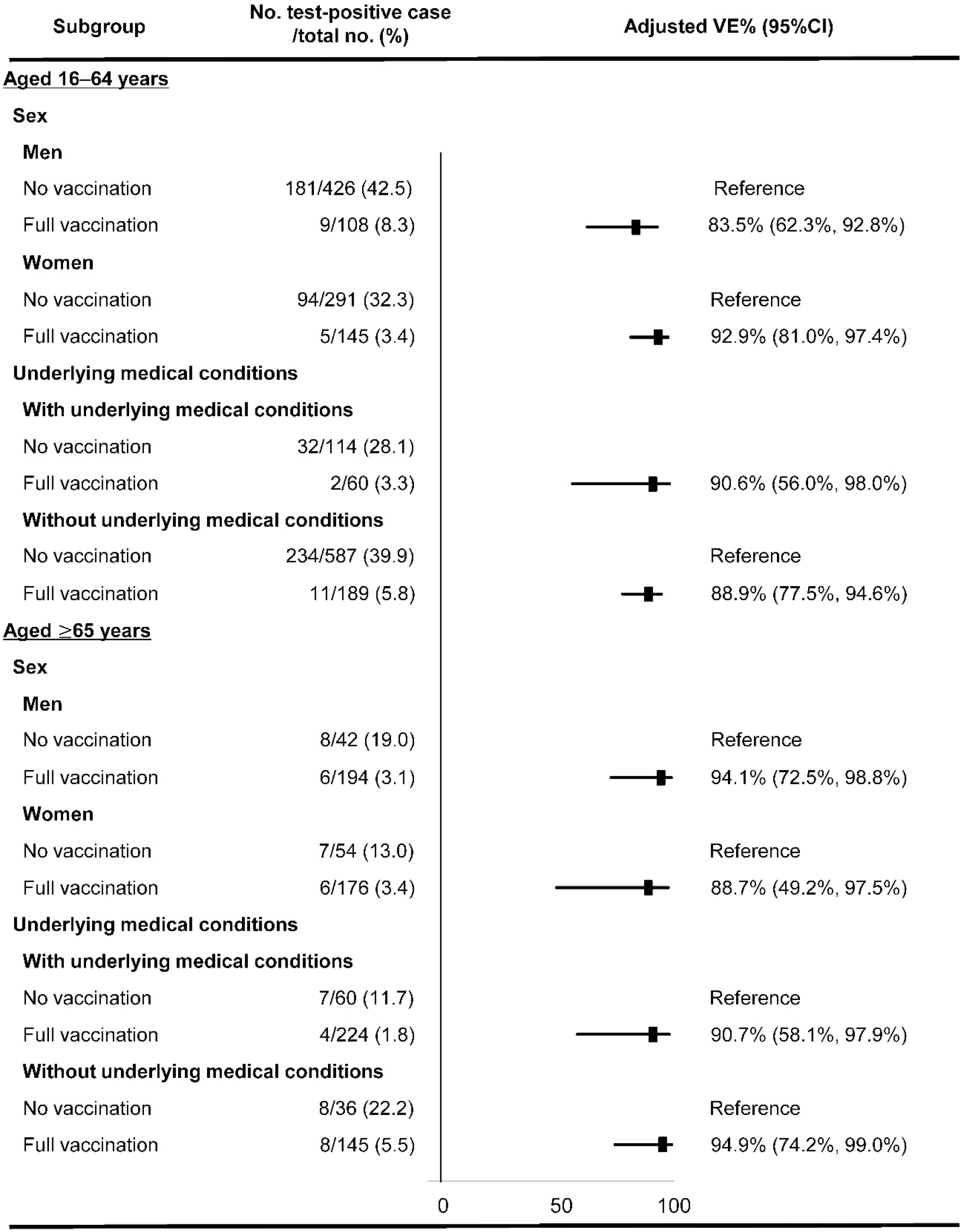
Vaccine effectiveness of messenger RNA COVID-19 vaccines against symptomatic SARS-CoV-2 infections by subgroups among individuals aged 16 to 64 years and aged ≥65 years, VERSUS study, Japan, July 1–September 30, 2021. The analysis included test-positive cases who had signs or symptoms comparable with COVID-19 and tested positive for SARS-CoV-2, and test-negative controls who had signs or symptoms comparable with COVID-19 and tested negative for SARS-CoV-2. Vaccine effectiveness was adjusted for age, sex, presence of underlying medical conditions, calendar week, history of contact with COVID-19 patients, and study site. Vaccination status was classified into three statuses based on the number of vaccine doses received before symptom onset and the number of days between the last vaccination date and symptom onset; no vaccination where individuals had received no vaccine dose before symptom onset; partial vaccination where individuals received one dose ≥14 days before symptom onset; and full vaccination where individuals who received two doses ≥14 days before symptom onset. Underlying medical conditions included chronic heart disease, chronic respiratory disease, obesity (body mass index≥ 30kg/m2), malignancy (including solid or haematological malignancy), diabetes, chronic kidney disease, receiving dialysis, liver cirrhosis, use of immunosuppressive medicines, or pregnancy. Abbreviations: VE, vaccine effectiveness; COVID-19, coronavirus disease 2019; SARS-CoV-2, severe acute respiratory syndrome coronavirus 2; VERSUS, Vaccine Effectiveness Real-time Surveillance for SARS-CoV-2.

## Discussion

In this prospective test-negative case-control study, we confirmed high mRNA COVID-19 vaccine effectiveness against symptomatic SARS-CoV-2 infections in Japan. We estimated that the vaccine effectiveness of two doses of BNT162b2 or mRNA-1273 against symptomatic SARS-CoV-2 infections was 88.7% (95% CI, 78.8–93.9) among patients aged 16 to 64 years and 90.3% (95% CI, 73.6–96.4) among patients aged ≥65 years. This study adds to real-world evaluations that demonstrated the high vaccine effectiveness of mRNA COVID-19 vaccines against symptomatic SAR-CoV-2 infections in Japan.

The patients included in this study were those examined for SARS-CoV-2 tests between July 1, 2021 and September 30, 2021. During this period, the Delta variant was dominant within Japan, and more than 90% of COVID-19 cases nationwide were estimated to be caused by the Delta variant since late August. [25, 26]. Therefore, both mRNA COVID-19 vaccines are effective against symptomatic SARS-CoV-2 infections caused by the Delta variant. Our vaccine effectiveness estimates after two doses of vaccines were similar to estimates reported from the United Kingdom (88% [95% CI, 85.3–90.1] for BNT162b2) [27] and Canada (92% [95% CI, 89–94] for BNT162b2 and 92% [95% CI, 90–97] for mRNA-1273) [28]; but higher than those in Israel (40.5% [95% CI, 8.7–61.2] for BNT162b2) [29, 30], the United States (42% [95% CI, 13–62] for BNT162b2 and 76% [95% CI, 58–87] for mRNA-1273) [31], or Qatar (44.4% [95% CI, 37.0–50.9] for BNT162b2 and 73.9 [95% CI, 65.9–79.9] for mRNA-1273) [32]. The Japanese national COVID-19 vaccination campaign started more than two months after these countries [33], and the symptom onset date for approximately 87% of the fully vaccinated patients in our study was one to three months after full vaccination. One reason for the difference between our estimates of vaccine effectiveness and those in Israel, the United States, and Qatar could be due to waning immunity [29, 34, 35]. On the other hand, the study in the UK included SARS-CoV-2 test results in late April to May [27]; whereas in Canada, the vaccination coverage rate started increasing since June 2021 [33] as in Japan (Supplementary Figure 2), which could make waning immunity less than in Israel, the United States, or Qatar [27, 28]. We also evaluated the vaccine effectiveness of each mRNA COVID-19 vaccine among people aged 16 to 64 years. Vaccine effectiveness of mRNA-1273 was higher than that of BNT162b2 in the point estimates, consistent with previous studies [31, 32]; however, there was no statistically significant difference (p–value=0.877).

As mentioned above, COVID-19 vaccinations were publicly funded in Japan, just after the market approval. This policy would be reasonable under the first phase of the pandemic situation. However, when we re-consider the national COVID-19 vaccination policy, such as a re-setting of priority-based booster vaccinations, subgroup analyses, such as stratified by age group, are needed. Older adults were reported to have lower antibody titers after COVID-19 vaccination compared to younger people [36-38]. On the other hand, COVID-19 vaccine effectiveness in older adults compared to younger people varied by studies. For example, in our study, the vaccine effectiveness of two doses of mRNA COVID-19 vaccines against symptomatic SARS-CoV-2 infections among patients aged ≥65 years was similar to that among patients aged 16 to 64 years, consistent with studies from Canada and Israel [28, 29]; whereas a study from the United Kingdom reported that vaccine effectiveness of BNT162b2 against symptomatic SARS-CoV-2 infections caused by the Delta variant was lower in patients aged ≥65 years than in patients aged 16 to 64 years [39]. Because COVID-19 vaccine effectiveness by age group differed by studies, it is crucial to continue assessing it.

To evaluate the waning immunity of mRNA COVID-19 vaccines against symptomatic COVID-19, we assessed vaccine effectiveness by dividing the interval between full vaccination date and symptom onset date into two groups. Although the 95% confidence interval was wide due to the small sample size, the point estimates were slightly lower in patients with a longer interval after vaccination. This result was consistent with studies in Israel, the United States, and the United Kingdom [29, 34, 39].

This study had several limitations. First, the sample size was limited to 13 study sites in nine prefectures between July 1, 2021, and September 30, 2021. Second, recall bias could occur in vaccination histories. In Japan, medical professionals working at medical institutions are not allowed to access governmental vaccination records, and vaccination histories included in this analysis were obtained through interviews with patients or their family members. To strengthen our results, we conducted several sensitivity analyses (Supplementary Table 1). Vaccine effectiveness obtained from sensitivity analyses was similar to the primary analysis, and we considered our results robust. Third, since we didn’t conduct SARS-CoV-2 genome sequencing for test-positive patients, it was impossible to obtain an accurate estimation of vaccine effectiveness of mRNA COVID-19 vaccines against the Delta variant. Fourth, we included test methods other than PCR when determining cases and controls because in Japan, test methods such as LAMP and antigen quantification have been used as commonly as PCR. Since these tests have acceptable levels of sensitivity and specificity, we thought it reasonable to include these test methods.

In conclusion, mRNA COVID-19 vaccines were highly effective for preventing symptomatic SARS-CoV-2 infections in Japan from July to September 2021, when the Delta variant circulated nationwide. Thus, vaccine effectiveness of mRNA COVID-19 vaccines remained high in Japan despite the dominancy of a variant virus. As we only evaluated the waning immunity up to six months after the full vaccination, and the sample size within three to six months after the full vaccination was limited, further follow-up research is needed.

Vaccination is one of the essential strategies to tackle the COVID-19 pandemic, and it is crucial to continue this surveillance activity, including the evaluation of vaccine effectiveness against the Omicron variant, to assess the optimal domestic COVID-19 vaccination strategy.

## Data Availability

All data produced in the present study are not available because the intellectual property agreement with the facilities has not been completed.

## Acknowledgements

We are grateful to the medical staff at the participating hospitals and clinics. For their assistance, we thank Yumi Araki, Kyoko Uchibori, Rina Shiramizu, Fumiyo Tsujita (Institute of Tropical Medicine, Nagasaki University), and Daichi Hatahara (Nagasaki University). We thank Yura Ko (National Institution of Infectious Diseases) for his technical support.

