## Supplementary Table 1 for "Effectiveness of mRNA COVID-19 vaccines against symptomatic SARS-CoV-2 infections during the Delta variant epidemic in Japan: Vaccine Effectiveness Real-time Surveillance for SARS-CoV-2 (VERSUS)"

**Supplementary Table 1:** Sensitivity analysis for vaccine effectiveness of mRNA COVID-19 vaccines against symptomatic SARS-CoV-2 infections: VERSUS study, Japan, July–September 2021

| | **Adjusted odds ratio (95% CI)** | **Vaccine effectiveness (%) (95% CI)** |
|---|---|---|
| **Full vaccination** | | |
| **Patients aged 16 to 64 years** | | |
| BNT162b2 or mRNA-1273 | | |
| No vaccination | 1.000 | |
| Scenario A | 0.116 (0.062 to 0.216) | 88.4 (78.4 to 93.8) |
| Scenario B | 0.112 (0.060 to 0.210) | 88.8 (79.0 to 94.0) |
| Scenario C | 0.093 (0.046 to 0.187) | 90.7 (81.3 to 95.4) |
| Scenario D | 0.114 (0.061 to 0.213) | 88.6 (78.7 to 93.9) |
| BNT162b2 | | |
| No vaccination | 1.000 | |
| Scenario A | 0.134 (0.068 to 0.267) | 86.6 (73.3 to 93.2) |
| Scenario B | 0.133 (0.067 to 0.263) | 86.7 (73.7 to 93.3) |
| Scenario C | 0.130 (0.064 to 0.264) | 87.0 (73.6 to 93.6) |
| mRNA-1273 | | |
| No vaccination | 1.000 | |
| Scenario A | 0.035 (0.044 to 0.278) | 96.5 (72.2 to 99.6) |
| Scenario B | 0.034 (0.004 to 0.269) | 96.6 (73.1 to 99.6) |
| Scenario C | N/A* | N/A* |
| **Patients aged ≥65 years** | | |
| BNT162b2 or mRNA-1273 | | |
| No vaccination | 1.000 | |
| Scenario A | 0.098 (0.036 to 0.267) | 90.2 (73.3 to 96.4) |
| Scenario B | 0.098 (0.036 to 0.266) | 90.2 (73.4 to 96.4) |
| Scenario C | 0.123 (0.045 to 0.333) | 87.7 (66.7 to 95.5) |
| Scenario D | 0.113 (0.042 to 0.303) | 88.7 (69.7 to 95.8) |
| BNT162b2 | | |
| No vaccination | 1.000 | |
| Scenario A | 0.142 (0.050 to 0.406) | 85.8 (59.4 to 95.0) |
| Scenario B | 0.142 (0.050 to 0.406) | 85.8 (59.4 to 95.0) |
| Scenario C | 0.155 (0.054 to 0.442) | 84.5 (55.8 to 94.6) |

Abbreviations: mRNA, messenger RNA; COVID-19, coronavirus disease 2019; SARS-CoV-2, severe acute respiratory syndrome coronavirus 2; VERSUS, Vaccine Effectiveness Real-

time Surveillance for SARS-CoV-2; CI, confidence interval.
Scenario A, for those without precise vaccination dates, we set the most recent possible dates from symptom onset as vaccination dates.
Scenario B, for those without precise vaccination dates, we set the earliest possible dates from symptom onset as vaccination date.
Scenario C, patients with no information of vaccination date were excluded.
Scenario D, multiple imputations were performed for those whose COVID-19 vaccination histories were not documented.
Full vaccination was defined as receiving two doses ≥14 days before the symptom onset.
*We could not evaluate the odds ratio because the odds ratio was tiny, almost zero.
