## Supplementary Figure 1 for "Effectiveness of mRNA COVID-19 vaccines against symptomatic SARS-CoV-2 infections during the Delta variant epidemic in Japan: Vaccine Effectiveness Real-time Surveillance for SARS-CoV-2 (VERSUS)"

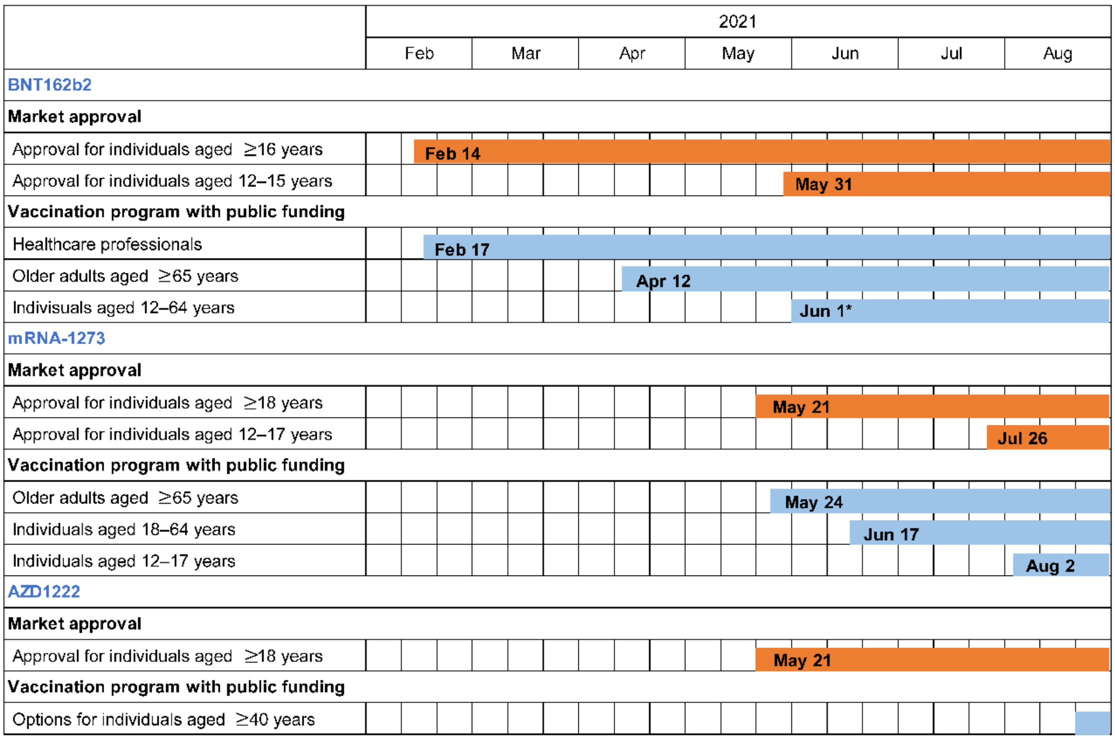


**Supplementary Figure 1**: Timeline of COVID-19 vaccine approval and administration in Japan. *Vaccination of individuals aged 12 to 64 years prioritized those with underlying medical conditions.

The data source was the National Institute of Infectious Diseases, Japan and the Pharmaceuticals and Medica Devices Agency, Japan.
