## Supplementary Figure 2 for "Effectiveness of mRNA COVID-19 vaccines against symptomatic SARS-CoV-2 infections during the Delta variant epidemic in Japan: Vaccine Effectiveness Real-time Surveillance for SARS-CoV-2 (VERSUS)"

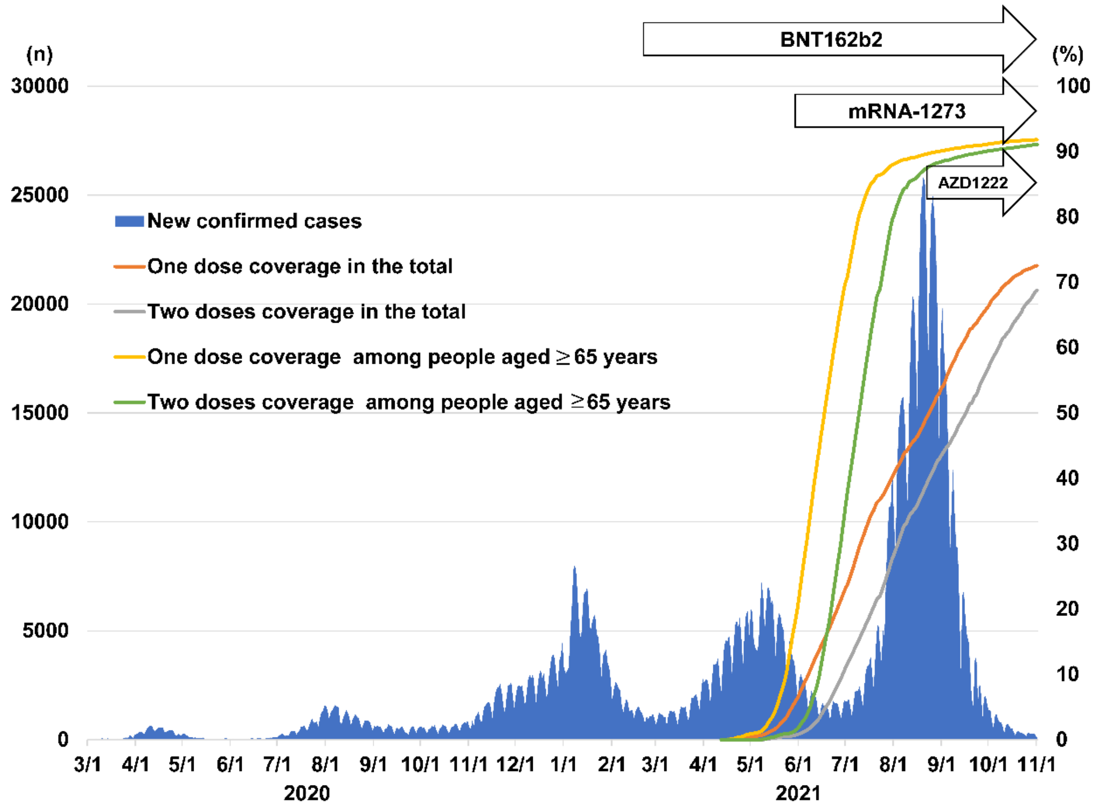


**Supplementary Figure 2:** The incidence of COVID-19 cases in Japan from March 1, 2020, through November 1, 2021. Japan has experienced five waves of COVID-19 epidemics. This study used registered data for surveillance between July 1, 2021, and September 30, 2021, during the fifth wave. Y-axis: the number of new COVID-19 cases per day. The orange, grey, yellow, and green lines indicate the proportion of persons vaccinated with one dose to Japan's total population, the proportion of persons vaccinated with two doses to Japan's total population, the proportion of one-dose vaccinated people aged ≥65 years to the total population aged ≥65 years, and the proportion of two doses vaccinated people aged ≥65 years to the total population aged ≥65 years, respectively.

The number of COVID-19 cases per day was obtained from the Ministry of Health, Labour and Welfare, Japan. The percentage of vaccinated persons was obtained from the Government CIOs' Portal, Japan.
