## Supplementary Figure 3 for "Effectiveness of mRNA COVID-19 vaccines against symptomatic SARS-CoV-2 infections during the Delta variant epidemic in Japan: Vaccine Effectiveness Real-time Surveillance for SARS-CoV-2 (VERSUS)"

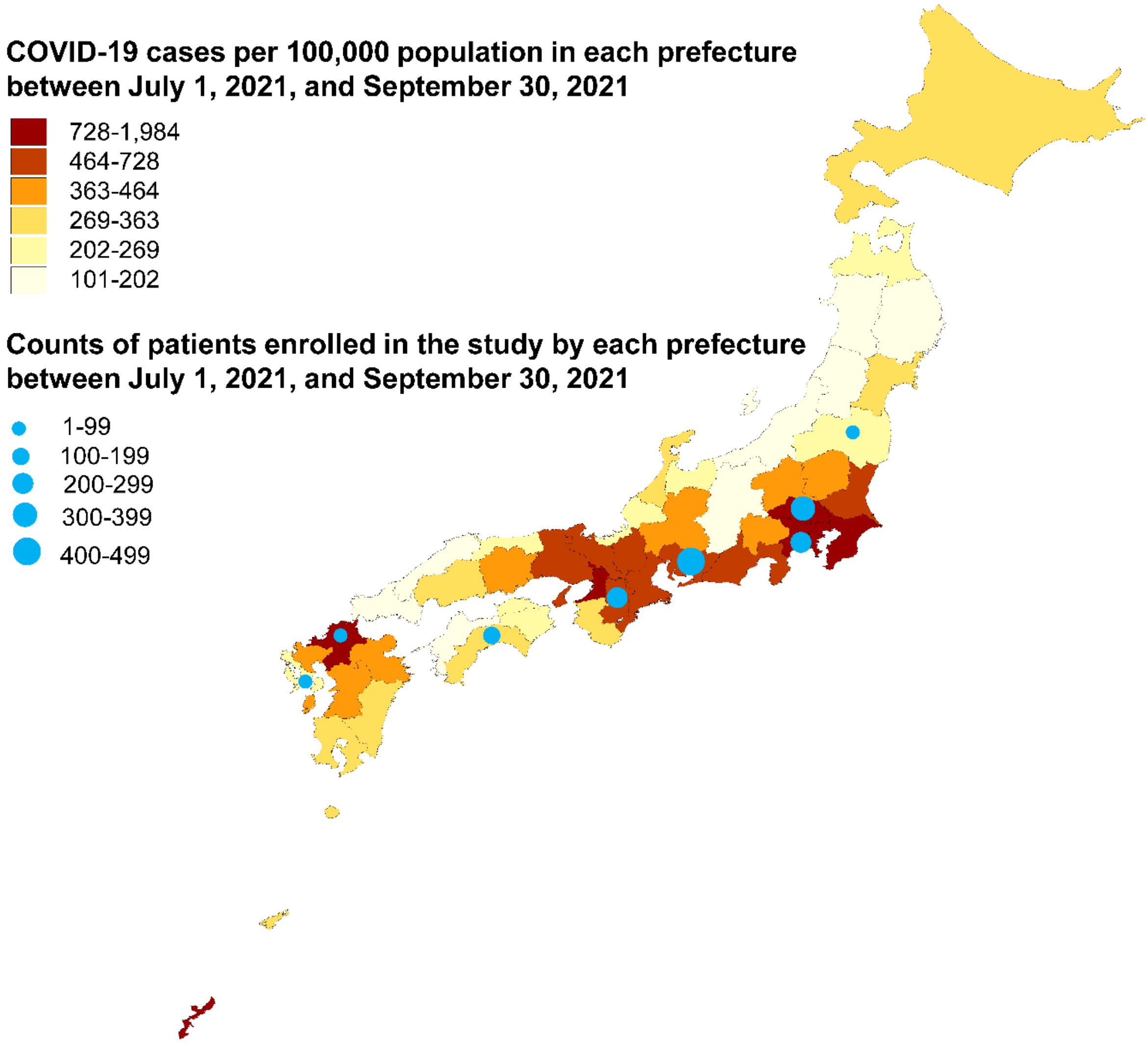


**Supplementary Figure 3:** Map of Japan with incidence of COVID-19 cases by prefecture between July 1, 2021, and September 30, 2021. Counts of patients enrolled in participating hospitals or clinics in each prefecture are shown on the map with circles. The size of each circle represents the number of patients included from each prefecture in this study.
Abbreviations: COVID-19, coronavirus disease 2019.
Data of COVID-19 cases per 100,000 in each prefecture between July 1, 2021, and September 30, 2021, was obtained from the Ministry of Health, Labour and Welfare, Japan.
